# Data-driven identification of unusual prescribing behaviour: an analysis and interactive data tool using six months of primary care data from 6500 practices in England

**DOI:** 10.1101/2022.09.22.22280200

**Authors:** Lisa EM Hopcroft, Jon Massey, Helen Curtis, Brian MacKenna, Richard Croker, Orla Macdonald, David Evans, Peter Inglesby, Seb Bacon, Tom O’Dwyer, Ben Goldacre, Alex J Walker

## Abstract

**Background:** Approaches to addressing unwarranted variation in healthcare service delivery have traditionally relied on the prospective identification of activities and outcomes, based on a hypothesis, with subsequent reporting against defined measures. Practice-level prescribing data in England are made publicly available by the NHS Business Services Authority for all general practices. There is an opportunity to adopt a more data-driven approach to capture variability and identify outliers by applying hypothesis free data driven algorithms to national datasets.

**Objectives:** To develop and apply a hypothesis free algorithm to identify unusual prescribing behaviour in primary care data at multiple administrative levels in the NHS in England, and to visualise these results using organisation-specific interactive dashboards.

**Methods:** Here we report a new data-driven approach to quantify how ‘unusual’ prescribing rates of a particular chemical within an organisation are as compared to peer organisations, over a period of six months (June-December 2021). This is followed by ranking to identify which chemicals are the most notable outliers in each organisation. These outlying chemicals are calculated for all practices, primary care networks, clinical commissioning groups and sustainability and transformation partnerships in England. Results are presented via organisation-specific interactive dashboards, the iterative development of which has been informed by user feedback.

**Results:** User feedback and internal review of case studies demonstrate that our methodology identifies chemicals that are in line with local policies and internal reporting.

**Conclusions:** Data-driven approaches overcome existing biases with regards to the planning and execution of audits, interventions and policy-making within NHS organisations, potentially revealing new targets for improved healthcare service delivery. We provide our dashboards as a candidate list for the consideration of expert users to prioritise for further interpretation and qualitative research in terms of potential targets for improved performance.

## Introduction

There is a recognition that evidence-based decision making in the NHS in England is critical to maintaining standards of care while reducing NHS spending [1] and the UK government has recently consulted on wide ranging plans to “digitise, connect and transform the health and care sector”, with a key priority being data-driven innovation. Flagship initiatives such as Getting it Right First Time [2] and RightCare [3] focus on identifying and addressing unwarranted variation in the NHS. Such initiatives can be limited in their scope in that the “data-driven” element of the work often focuses on assessing performance relative to recommendations that are defined prospectively, rather than employing hypothesis free data-driven methodologies to make objective assessments as to where opportunities for improvement might exist.

Monthly prescription data for every general practice in England has been made available to the public since 2010 [4]. This dataset includes product and month of prescribing, the number of items prescribed and the total quantity, making it very amenable to detailed analysis for the purposes of original research [5–9] and systematic audits and reviews [10,11]. Mining these data for unusual prescribing behaviour could identify where service delivery improvements could be made and help inform local decision makers when designing appropriate interventions and policies.

We run OpenPrescribing [12], a website that allows public interrogation and visualisation of these primary care prescription data at multiple administrative levels in the NHS in England. We have previously deployed novel methodologies to identify changes over time in any one of the 80 measures implemented in OpenPrescribing, providing monthly alerts to notify practitioners when their prescribing rates deviate from the norm and may require clinician attention [13]. These measures have been selected on the basis of clear guidance being available from health authorities and are subject to initial and continuing review by clinicians, pharmacists and epidemiologists.

We set out to develop new hypothesis-blind data science techniques to identify unusual prescribing behaviour and therefore potential opportunities for service improvement. We applied this methodology to the national prescribing dataset to identify outliers at multiple administrative levels of the NHS in England, presenting the most extreme outliers in each organisation for the consideration of expert users to prioritise for further review, qualitative research and interpretation within the local context.

## Methods

### BOX 1

**NHS England administrative organisations**

Primary care in England is delivered by individual general **practices**, with one or more general practitioners (GPs). Almost all (>99%) practices are grouped together with other local primary care provision to form **primary care networks (PCNs)**, typically representing 30,000-50,000 people [14]. **Clinical commissioning groups (CCGs)** are clinically-led organisations that are responsible for the commission of primary (and secondary) care in a geographical region [15]. As of April 2021, there were 106 CCGs in England. Each practice will belong to one CCG. Finally, CCGs are clustered into **Sustainability and Transformation Partnerships (STPs)** [16]. As of May 2020, there were 42 STPs in England. In July 2022, CCGs and STPs were replaced with 42 Integrated Care Boards (ICBs) though the data used in this study predates this change.

### Study design

Prescribing practice was analysed by conducting a retrospective cohort study using prescribing data from all English NHS general practices, PCNs, CCGs and STPs.

### Data Source

Data for the period June 2021 to December 2021 were extracted from the OpenPrescribing database. This imports openly accessible prescribing data from the large, monthly files published by the NHS Business Services Authority (BSA), which contain data on cost and items prescribed for each month for every typical general practice and CCG in England, dating back to mid-2010 [4]. Detailed methods for the creation of OpenPrescribing, including data management, aggregation and cleaning, are available elsewhere [17]. The monthly prescribing datasets contain one row for each different medication and dose in each prescribing organisation in NHS primary care in England, describing the number of items (that is, prescriptions issued) and the total cost. These data are sourced from community pharmacy claims data and, therefore, contain all items that were dispensed. All available prescribing data were extracted for institutions identified as typical general practices; all other organisations, such as prisons or specialist community clinics, were excluded using NHS Digital organisation data [18]. We limited our analysis to the 2369 chemicals from chapters 1-15 of the BNF, to exclude chapters not following a chemical/sub-paragraph structure, those which largely cover non-medicinal products such as dressings.

### Outlier detection

We were interested in detecting outliers with regards to *chemicals* (so as to combine all presentations of the same chemical in one item). We first calculate a prescription rate for each chemical in each practice; specifically, we calculate the number of prescriptions containing our chemical of interest, and divide this by the number of prescriptions containing chemicals of the same BNF subparagraph, for example all statin prescriptions as a proportion of all lipid regulating drugs. This captures the prescribing rate for the chemical of interest as compared to all drugs in the same class in a single practice This ratio is calculated across all practices and the mean and standard deviation calculated. The ratios in each practice are then re-expressed as Z scores using this mean and standard deviation. A Z score is the number of standard deviations that a given data point is away from the mean. The Z scores are used to rank all chemicals within a practice in terms of their outlier status (the most extreme outliers occupying the top and bottom of this ranked list).

This process is repeated at three higher administrative levels—PCN, STP and CCG—to generate the equivalent ranked list of prescribing outliers for these larger organisations.

### Organisation-level results visualisation

An interactive dashboard has been created at OpenPrescribing.net [19] for each organisation where data describing twenty of the most extreme outliers is summarised: ten where prescribing in the organisation is *higher* than other peer organisations, and ten where prescribing in the organisation is lower than other peer organisations. Tables are provided for both sets, which summarise the various values described in the Methods: *Chemical Items* and *Subparagraph Items* are the number of prescriptions for the chemical and BNF subparagraph respectively; Ratio is the *Chemical Items* as a proportion of *Subparagraph Items* for the chemical in the organisation of interest; *Mean* and *Std* summarise this ratio over all organisations and *Z_score* is the *Ratio* re-expressed as a Z score. This same information is described visually by a density plot, where the distribution of ratios across all organisations is captured by a blue line, with the ratio for the organisation of interest indicated by a vertical red line. Densities are generated using Seaborn’s kdeplot() function, setting the bandwidth for smoothing as suggested by [20].

### User feedback

Early prototypes were shared directly with interested parties and any feedback gained was used to inform iterative development of the tool and proposed visualisations of the results. Further to this, the tool was shared more widely (via Twitter), and formal feedback was collected via a Google form (Box 2). Additional unstructured feedback was compiled from direct emails and mentions on social media.

#### BOX 2

**Outlier detection feedback form**

**Respondent details**

*Email* Free text

*Which organisation’s report are you giving feedback on?* Free text

*Please describe your relationship to the organisation (e*.*g. doctor, practice nurse, commissioner)* Free text

**Understandability**

*Does this report make sense to you?* Yes/No.

*Any further comments on the understandability of the report(s)* Free text

**Interest**

*Is it interesting?* Yes/No.

*Any further comments on the interestingness of the report(s)* Free text

**Utility**

*Is it useful?* Yes/No.

*Any further comments on the usefulness of the report(s)* Free text

**Individual items**

*Thinking about where your prescribing is higher than most, please describe any observations you have on any individual items*. Free text

*Thinking about where your prescribing is lower than most, please describe any observations you have on any individual items*. Free text

**Improvements**

*What, if anything, would you change about the report(s)?* Free text

### Software and Reproducibility

Data management was performed using Python 3.8.1 and Google BigQuery, with analysis carried out using Python. Code for data management and analysis are archived online [21] and dashboards are available on the OpenPrescribing website [19].

### Patient and Public Involvement

We publicised this tool via social media and actively sought feedback from interested healthcare professionals and members of the public to inform iterative development via a survey (see Feedback section above). We will continue to seek and consider feedback via these same channels as the tool is developed. We have developed a publicly available website [12] through which we invite any patient or member of the public to contact us regarding this study or the broader OpenPrescribing project.

## Results

### Outlier detection

Summary statistics for Z scores calculated for all chemicals across each type of organisation are shown in the “All chemicals” portion of Table 1. A median of 0.00 and narrow IQRs across all organisations demonstrate that the prescribing rates for the majority of chemicals is consistent across peer organisations. The maximum and minimum values increase and decrease respectively with the size of the organisation: the most extreme outliers occur further away from the mean as the organisation size decreases.

**Table 1:**
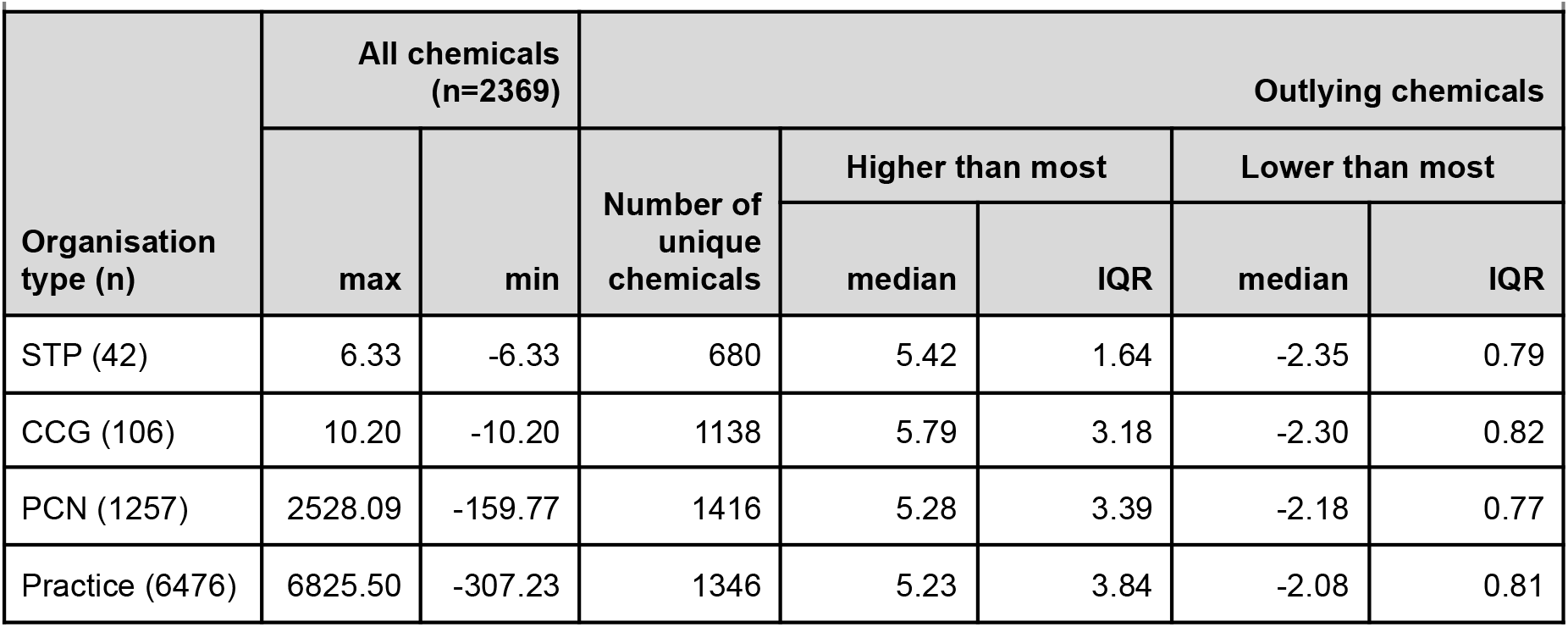
Summary statistics for Z scores calculated for All chemicals (n=2369) and Outlying chemicals, for all Organisation Types (number of organisations of this type provided in brackets). Outlying chemicals are those that occur in the top ten (i.e., ‘Higher than most’) or bottom ten (i.e., ‘Lower than most’) by Z score, in at least one organisation of the relevant type.

Summary statistics for Z scores of outlying chemicals (i.e., the ten chemicals ranked highest and ten chemicals ranked lowest by Z score) across each type of organisation are shown in the “Outlying chemicals” portion of Table 2. While the median values for the ‘higher than most’ outlying chemicals are similar, the IQR values demonstrate that variation between peer organisations decreases with the size of the organisation: the least amount of variation is observed between STPs and the most amount of variation is observed between practices. More outlying chemicals are identified in smaller organisations (PCNs and practices). With regards to outlying chemicals identified as being prescribed at lower rates compared to peer organisations, both the median and IQR of the Z scores is very similar across all organisation types. The Z scores for higher than most outlying chemicals are more extreme than the lower than most outlying chemicals in all organisation types.

### Organisation-level results visualisation

An example dashboard showing the outlying chemicals in one STP[22] is shown in Figure 1. The first table lists the top ten chemicals that are prescribed at *higher* rates here compared to other STPs; the second table lists the top ten chemicals that are prescribed at *lower* rates here compared to other STPs. Focussing on the results for co-codamol (in the second, “lower than most” table), we can see that 20% (270,523) of the 1,353,879 “Non-opioid analgesics and compound preparations” items contain co-codamol and that this is 3 standard deviations (SDs) below the mean for all STPs. The sparkline plot provided demonstrates visually where this 20% falls (red line) in the distribution across all STPs (blue line).

### Case study: NHS Devon CCG

NHS Devon CCG is the fifth largest CCG in England, commissioning healthcare for 1.2 million people in the South West of England. The prescribing outliers for this STP as identified by our tool are shown in Figure 2 and have been reviewed by the local medicines optimisation team who provide likely explanations for outlier prescribing.

Several of the chemicals prescribed more often in NHS Devon CCGs than other CCGs are defined as first-line treatments in local formularies, for example flumetasone pivalate [23] and levofloxacin [24]. Corresponding patterns of under-prescribing can be see in the “lower than most” results table for similar chemicals, specifically ciprofloxacin (an alternative to levofloxacin) and dexamethasone (an alternative to flumetasone pivalate).

The lower prescribing rates for fusidic acid reflect a change in this CCG to prescribe this chemical by specialist recommendation only [25], due to rising costs [26] and a narrow spectrum of action. The lower rates of prescribing for senna and lactulose are also likely due to a formulary shift in this CCG towards macrogols [27]. Finally, the low prescribing rate of betamethasone esters is also expected as these chemicals are non-formulary in this CCG [28].

This dashboard also demonstrates a valid use for the low number results. Gluten free pastas and cereals are not available on the NHS, so should not appear at all. The identification of this low number outlier via our methodology has prompted further work within NHS Devon CCG to clarify how this prescription was generated and processed.

**Figure 1a:**
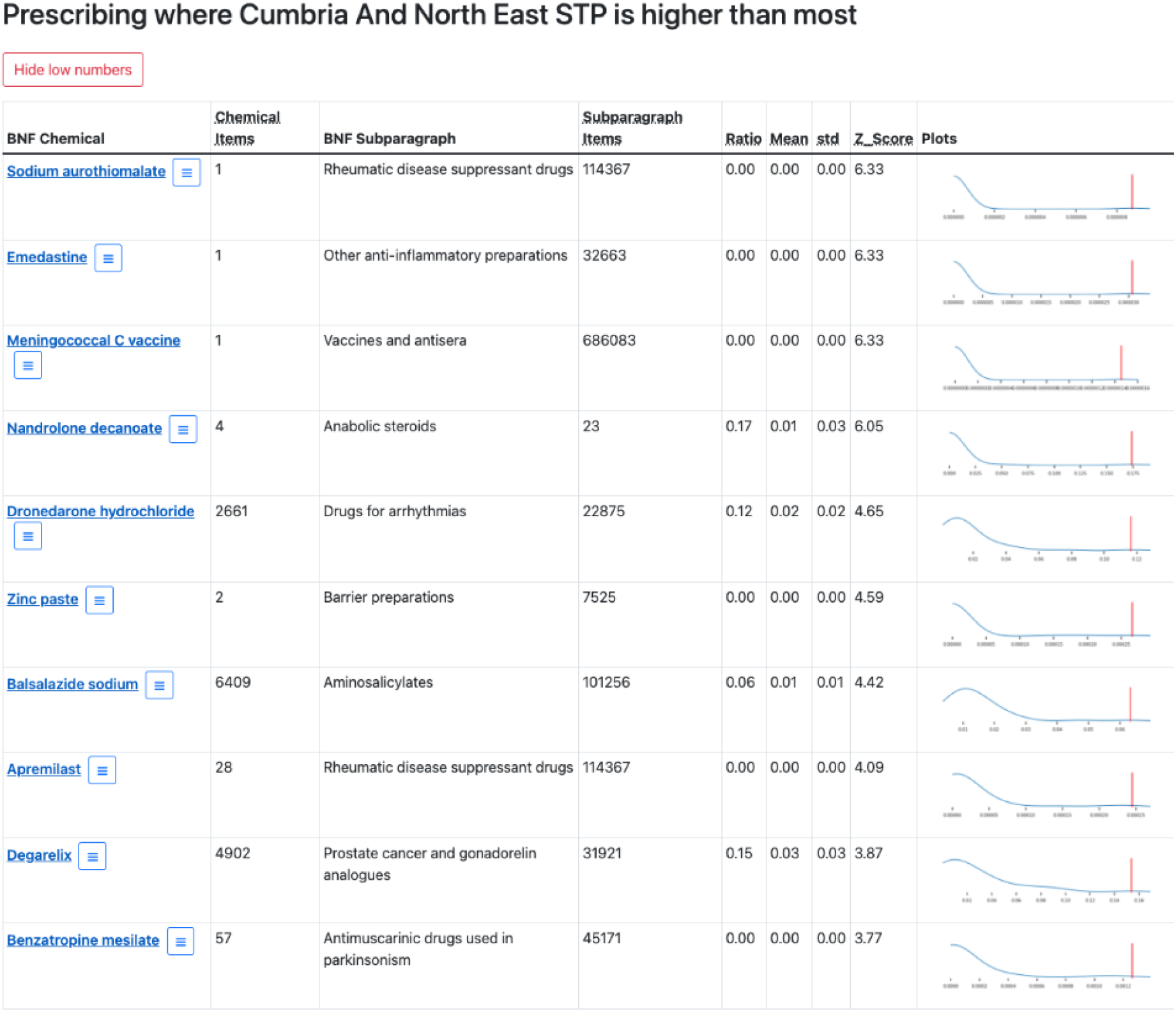
Example dashboard showing the top ten outlying chemicals for Cumbria and North East STP. BNF Chemical is the chemical of interest, Chemical Items provides the number of prescribing items containing this chemical. BNF Subparagraph is the BNF Subparagraph to which the Chemical belongs and Subparagraph Items is the number of prescribing items containing an item belonging to this BNF Subparagraph. Ratio, Mean, std and Z-score place the chemical items count in the context of the subparagraph items count as described in the methods. The sparkline plot shows where the Ratio value for this STP occurs (vertical red line) in the context of the same Ratio in all STPs (summarised by the blue line).

**Figure 1b:**
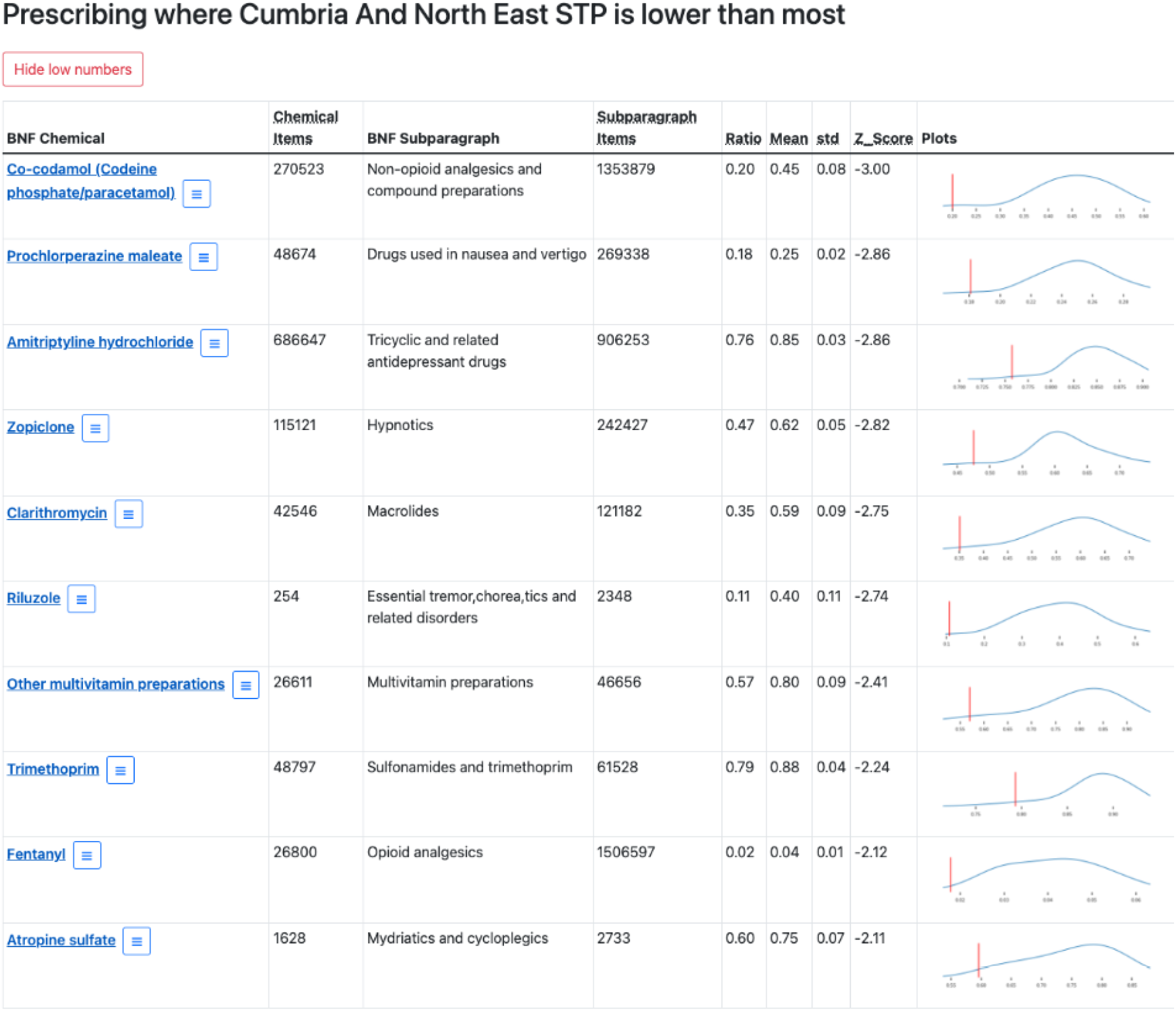
Example dashboard showing the bottom ten outlying chemicals for Cumbria and North East STP. See Figure 1a for definitions for each column.

**Figure 2a:**
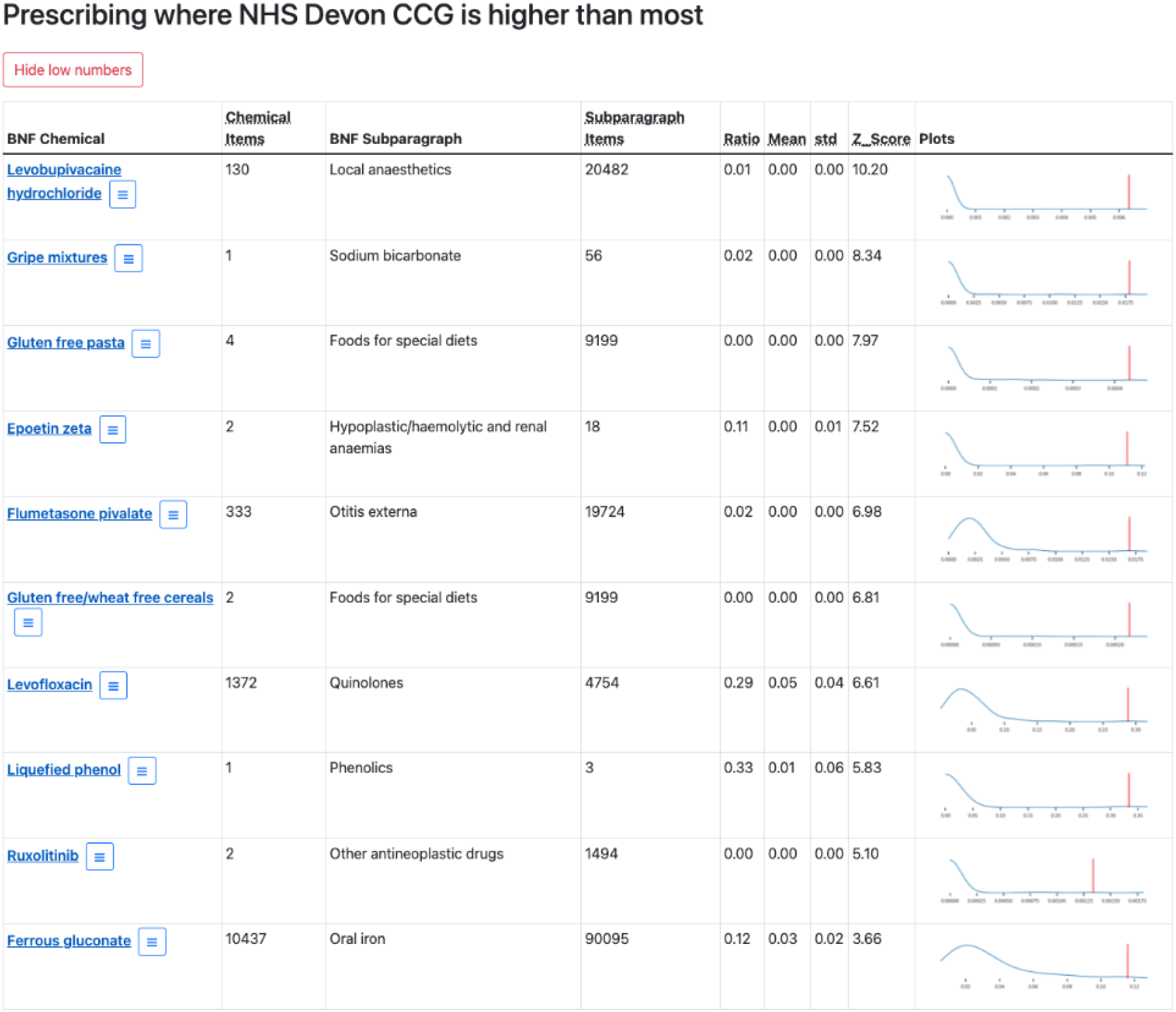
The top ten outlying chemicals for Devon CCG. See Figure 1a for definitions for each column.

**Figure 2b:**
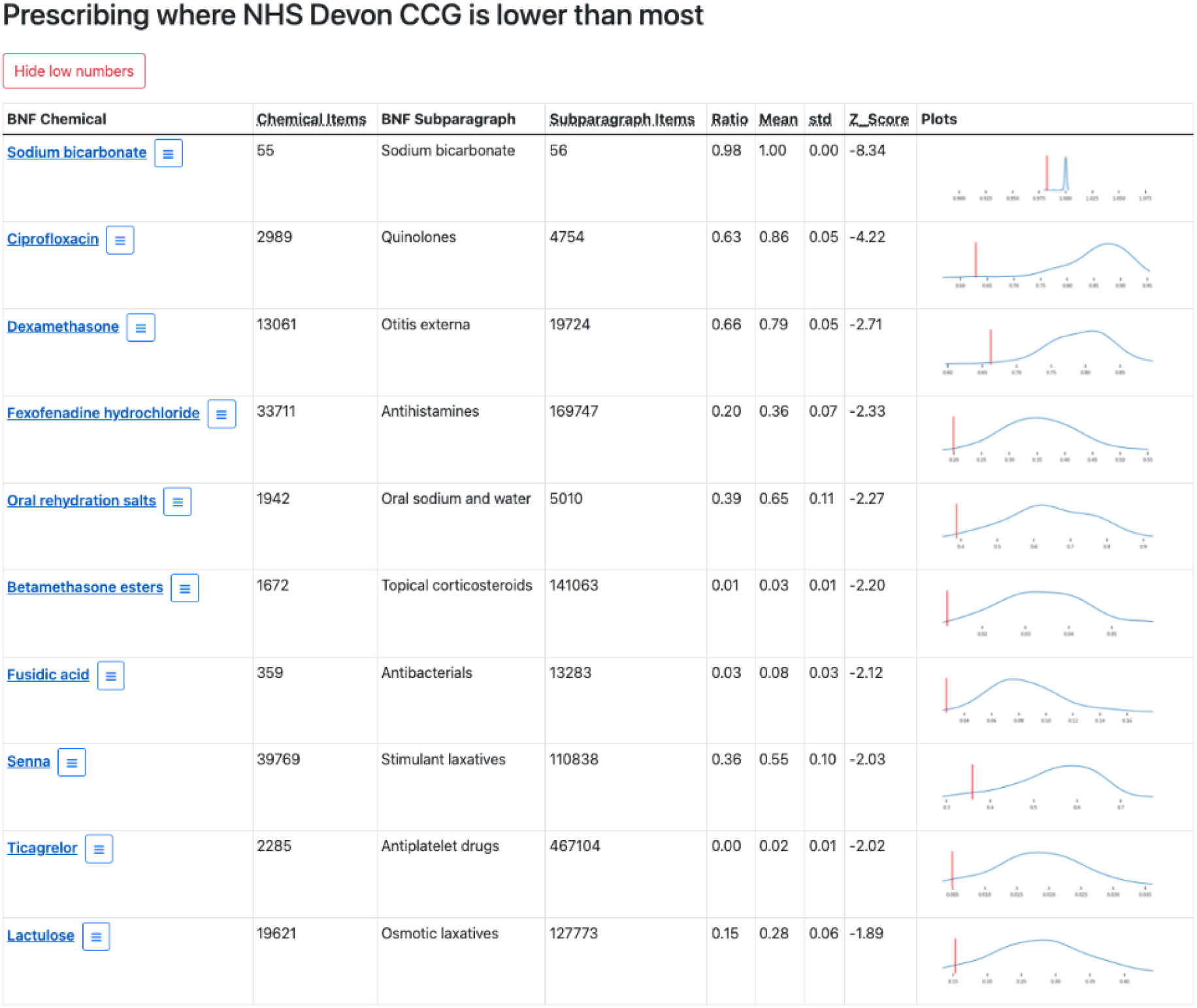
The bottom ten outlying chemicals for Devon CCG. See Figure 1a for definitions for each column.

### User feedback

Through the formal Google form and direct correspondence with interested parties, we received feedback for a prototype version of the dashboard from six individuals. An example of this prototype is shown in Supplementary Figure 1, showing five top/bottom outlying chemicals. Several respondents indicated that the results were expected (i.e., results echoed internal reporting or were aligned with local prescribing policies); while this indicates that our tool is working, one user did question what the added value was above existing reporting. Other users stated that the tool had revealed unexpected results, worthy of follow-up.

There were multiple requests to present more than the top and bottom five results (e.g., the top and bottom 10 or 20 results) to explore the data in more detail. Users recognised that extreme outliers could be derived from very small numbers of patients or items; some requested that results with small counts be removed though others recognised that these may be important, particularly in practices or PCNs. There was a suggestion that users could choose to have low numbers suppressed or displayed, depending on whether their focus was systemic anomalies or rogue prescriptions. There were also requests to include other data in the results, including cost and highlighting drugs on the “*Not Suitable to Prescribe*” list.

There were other requests that are more relevant to the design of the tool, rather than the analysis itself. The feedback demonstrated that users required more information to interpret and understand the data (i.e., Z scores, ratios, means, standard deviations) and that with this additional explanation, more could be made of the graphical summary. There was also a request for an improved user experience regarding navigating to practices via the drop down sections (which could be implemented as an organisational search).

We used the most common feedback to inform further development and the released version of the dashboards now includes the top/bottom ten outlying chemicals and optional filtering of low numbers (Figure 1).

## Discussion

### Summary

We have developed and implemented a new hypothesis free methodology to detect unusual or “outlier” prescribing rates of chemicals in one organisation, in relation to all “peer” organisations. We have applied this methodology to a national dataset to quantify how typical the prescribing is for individual chemicals, at multiple administrative levels (practice, PCN, CCG and STP). We have sought and will continue to seek user feedback to inform development and incrementally improve usability and functionality.

Summary statistics demonstrate that prescribing amongst peer organisations is generally concordant and that variation increases as the size of the organisation decreases. The data also demonstrate however that outliers do occur at all administrative levels: while there is less variation between STPs, the median rate of over-prescribed and under-prescribed outliers amongst STPs is 6.19 and −2.77 respectively, both substantial differences from the mean. Ranking of these quantifications allows us to identify the most extreme outliers in terms of prescribing behaviour, at each organisational level. A case study of an individual CCG (NHS Devon) demonstrated that our methodology identified prescribing patterns that aligned with local prescribing guidance, but also detected patterns that warranted further investigation. It is not appropriate to formally assess the utility of our methodology as there are many legitimate reasons that a chemical may be an outlier in a particular organisation: prescribing guidance as defined by local formulary may differ from elsewhere; local prescribing policy may place responsibility for prescribing particular drugs in secondary care rather than primary care; clinicians may be reluctant to change medication for patients who are stable on a long established medication regime (in particular the elderly or vulnerable); or there is justified preference for other drugs in the same class. Given the complexities of interpreting these data, we present this tool as a starting point for NHS organisations to perform and plan internal audits rather than a definitive reporting tool.

### Strengths and weaknesses

Our approach combines a comprehensive national prescribing dataset with a well understood system for drug classification, thereby capturing the national context at high resolution and allowing the interpretation of prescribing behaviour for *all* chemicals in multiple administrative levels of the NHS in England. The methods employed are well established and easy to understand; readily amenable to visual presentation as graphs; and allow prioritisation of results by ranking. Our approach has utility in other contexts and repurposing to gain greater understanding of other NHS data (e.g., hospital prescriptions) would be straightforward.

We also note some limitations. Firstly, the calculation of Z scores using mean and standard deviation assumes a normal distribution. This is more likely to be the case where numbers of items prescribed are high (aggregated to STP or CCG) but may not be the case where number of items are low (aggregated to PCN or practices, or where the items are more rarely prescribed). Secondly, this approach can generate very large Z scores where standard deviations are tight or item numbers generally are very low. An example of this can be seen in Figure 1: while the ratio generated by the number of prescribed items containing Sodium aurothiomalate is very low (1/114367 = 8.74×10^−6^), the tight standard deviations observed across the whole population of STPs translates this small value into a large Z score. Expert users may be seeking out such results to identify very rare prescribing items (low number results did prove important in the NHS Devon case study), but they may also wish to suppress such results to focus on more commonly prescribed chemicals. To accommodate this, and in line with our user feedback, we have implemented the option to show or hide counts of five or less.

### Findings in Context

This is one of a suite of tools that we are seeking to develop at OpenPrescribing.net, each of which capture variability with a view to leveraging further insight from the datasets to which we have access. We make extensive use of decile plots to place individual organisations into a wider context [5,6,29] and have applied algorithms to identify when those individual organisations start to deviate from the rest of their peers [13]. We have also used deciles to summarise financial data and estimate potential savings if PPU costs were aligned with the lowest decile [7]. These methodologies all have the potential to support NHS organisations in England to guide audits, prioritise and shape new policies and, crucially, assess the impact of those interventions with regards to patient care and cost savings.

### Policy Implications and Interpretation

The Department of Health and Social Care consultation explicitly recognises the value of near real-time data release and the potential of data-driven insights to guide targeted policy making [30]. The methodology described here contributes towards that key priority by exposing specific patterns in data that warrant attention that may have otherwise been obscured. We do not advocate that our approach be used in isolation, but rather as a starting point for expert users to interpret within the local context and make evidence-based decisions about priorities and planning. By updating these dashboards on a regular basis, we hope to provide decision-makers with near real-time feedback so as to monitor performance and respond quickly when necessary.

### Future Research

Areas for further research include implementing monthly updates as part of OpenPrescribing; enhancing results output as determined by ongoing user feedback (e.g., new functionality, information or visualisations); and, as we have implemented for our CUSUM results, organisation specific alerts to notify staff where prescribing behaviour appears to be different to their peers.

### Summary

Capturing the variability in prescribing rates amongst peer organisations permits the hypothesis-free identification of prescribing outliers. We have applied such an analysis to a national prescribing dataset and made the most extreme prescribing outliers in each organisation publicly available as interactive dashboards. We intend that these dashboards prompt further qualitative analysis within the individual organisations to identify where service delivery improvements could be made.

## Supporting information

RECORD checklist

## Data Availability

All data used in the present study are publicly available at https://www.nhsbsa.nhs.uk/prescription-data/prescribing-data/english-prescribing-data-epd. Code for data management and analysis are archived online at https://github.com/ebmdatalab/openprescribing/blob/main/openprescribing/pipeline/management/commands/outlier_reports.py. The generated dashboards are available at https://openprescribing.net/labs/outlier_reports/.

https://github.com/ebmdatalab/openprescribing/blob/main/openprescribing/pipeline/management/commands/outlier_reports.py

https://openprescribing.net/labs/outlier_reports/

## Administrative

## Acknowledgements

We are grateful to wider NHS colleagues for discussions that have informed our work on this topic.

## Guarantor

BG is guarantor.

## Contributorship

**Conceptualization:** H.C., B.M., B.G., and A.J.W.

**Data curation:** J.M., D.E., P.I., and S.B.

**Formal analysis:** L.E.M.H., J.M., B.M., and A.J.W.

**Funding acquisition:** H.C., B.G., and A.J.W.

**Investigation:** L.E.M.H., J.M., H.C., B.M., R.C., O.M., and A.J.W.

**Methodology:** L.E.M.H., J.M., H.C., B.M., R.C., and A.J.W.

**Resources:** D.E., P.I., S.B., and T.O.D.

**Software:** J.M., D.E., P.I., S.B., and T.O.D.

**Supervision:** B.G.

**Visualization:** L.E.M.H., J.M., H.C., B.M., and A.J.W.

**Writing - original draft:** L.E.M.H., J.M., H.C., B.M., and A.J.W.

**Writing - review & editing:** L.E.M.H., J.M., H.C., B.M., R.C., O.M., B.G., and A.J.W.

## Abbreviations

BNF: British National Formulary
CCG: Clinical Commissioning Group
GP: General Practitioner
ICB: Integrated Care Board
NHS: National Health Service
PCN: Primary Care Network
SD: standard deviation
STP: Sustainability and Transformation Partnership

