## Supplementary Figure 1 for "Data-driven identification of unusual prescribing behaviour: an analysis and interactive data tool using six months of primary care data from 6500 practices in England"

### Prescribing where Cumbria And North East STP is higher than most

| BNF Chemical | Chemical Items | BNF Subparagraph | Subparagraph Items | Ratio | Mean | std | Z_Score | Plots |
|---|---|---|---|---|---|---|---|---|
| Cholesterol/simvastatin | 1 | Preparations for psoriasis | 34553 | 0.00 | 0.0 | 0.0 | 6.33 | 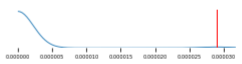 |
| Procarbazine hydrochloride | 1 | Other antineoplastic drugs | 1374 | 0.00 | 0.0 | 0.0 | 6.33 | 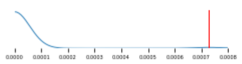 |
| Ephedrine hydrochloride | 1 | Vasoconstrictor sympathomimetics | 3356 | 0.00 | 0.0 | 0.0 | 6.33 | 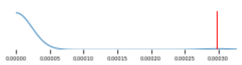 |
| Zidovudine | 1 | HIV infection | 37 | 0.03 | 0.0 | 0.0 | 6.25 | 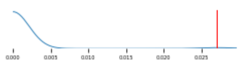 |
| Other topical circulatory preparations | 3 | Topical circulatory preparations | 1470 | 0.00 | 0.0 | 0.0 | 6.08 | 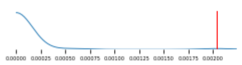 |

### Prescribing where Cumbria And North East STP is lower than most

| BNF Chemical | Chemical Items | BNF Subparagraph | Subparagraph Items | Ratio | Mean | std | Z_Score | Plots |
|---|---|---|---|---|---|---|---|---|
| Midodrine hydrochloride | 3355 | Vasoconstrictor sympathomimetics | 3356 | 1.00 | 1.00 | 0.00 | -6.33 | 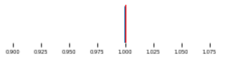 |
| Heparinoid | 1467 | Topical circulatory preparations | 1470 | 1.00 | 1.00 | 0.00 | -6.08 | 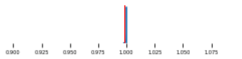 |
| Co-codamol (Codeine phosphate/paracetamol) | 193620 | Non-opioid analgesics and compound preparations | 963640 | 0.20 | 0.45 | 0.08 | -3.01 | 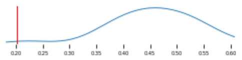 |
| Prochlorperazine maleate | 34255 | Drugs used in nausea and vertigo | 188214 | 0.18 | 0.25 | 0.02 | -2.84 | 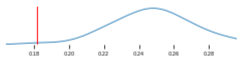 |
| Amitriptyline hydrochloride | 483140 | Tricyclic and related antidepressant drugs | 637512 | 0.76 | 0.85 | 0.03 | -2.83 | 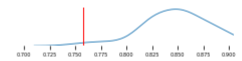 |

**Supplementary Figure 1: Prototype dashboard showing the top/bottom five outlying chemicals for Cumbria and North East STP.** BNF Chemical is the chemical of interest, Chemical Items provides the number of prescribing items containing this chemical. BNF Subparagraph is the BNF Subparagraph to which the Chemical belongs and Subparagraph Items is the number of prescribing items containing an item belonging to this BNF Subparagraph. Ratio, Mean, std and Z-score place the chemical items count in the context of the subparagraph items count as described in the methods. The sparkline plot shows where the Ratio value for this STP occurs (vertical red line) in the context of the same Ratio in all STPs (summarised by the blue line).
